# Identifying patients at high risk of inappropriate drug dosing in periods with renal dysfunction

**DOI:** 10.1101/2021.07.09.21257018

**Authors:** Benjamin Skov Kaas-Hansen, Cristina Leal Rodríguez, Davide Placido, Hans-Christian Thorsen-Meyer, Anna Pors Nielsen, Nicolas Dérian, Søren Brunak, Stig Ejdrup Andersen

## Abstract

**Introduction:** Dosing of renally cleared drugs in patients with kidney failure often deviates from clinical guidelines but little is known about what is predictive of receiving inappropriate doses.

**Methods and materials:** We combined data from the Danish National Patient Register and in-hospital data on drug administrations and estimated glomerular filtration rates for admissions between 1 October 2009 and 1 June 2016, from a pool of about 2.9 million persons. We trained artificial neural network and linear logistic ridge regression models to predict the risk of five outcomes (>0, ≥1, ≥2, ≥3 and ≥5 inappropriate doses daily) with index set 24 hours after admission. We used time-series validation for evaluating discrimination, calibration, clinical utility and explanations.

**Results:** Of 52,451 admissions included, 42,250 (81%) were used for model development. The median age was 77 years; 50% of admissions were of women. ≥5 drugs were used between admission start and index in 23,124 admissions (44%); the most common drug classes were analgesics, systemic antibacterials, diuretics, antithrombotics, and antacids. The neural network models had better discriminative power (all AUROCs between 0.77 and 0.81) and were better calibrated than their linear counterparts. The main prediction drivers were use of anti-inflammatory, antidiabetic and anti-Parkison’s drugs as well as having a diagnosis of chronic kidney failure. Sex and age affected predictions but slightly.

**Conclusion:** Our models can flag patients at high risk of receiving at least one inappropriate dose daily in a controlled in-silico setting. A prospective clinical study may confirm this holds in real-life settings and translates into benefits in hard endpoints.

## Introduction

Renal diseases affect patients’ susceptibility to, and modify the effects of many drugs, and they reduce renal clearance exposing patients to higher steady-state concentrations when given standard doses. The kidneys excrete active forms and/or metabolites of many drugs, so renal dysfunction necessitates dose-adjustment of renally cleared drugs with narrow therapeutic indices to prevent adverse events and accidental over-dosing.

Inadequate dose-adjustment of such drugs has been linked to polypharmacy [1,2] and can cause noxious events [3] or accidental over-dosing [4]. Although not a new issue, [5,6] deviating from guidelines is widespread with prevalence estimates up to 70% [1,2,7-9]. Despite large inter-individual variability in clearance and response, dose adjustment for many drugs is crude and based on the estimated glomerular filtration rate (eGFR), for example, halving the dose when eGFR < 60 ml/min/1.73 m^2^.

Appropriate alerts in order-entry systems may facilitate rational clinical decision-making, [10,11] and convincing examples have showcased how computerised systems can underpin rational pharmacotherapy [4,12]. However, downsides of extensive computerisation of healthcare emerge [13]; alert fatigue [14] is particularly problematic, and strategies and interventions have been proposed to mitigate its negative effects [15].

At Danish hospitals, prescriptions are mostly dispensed and administered by nurses who record detailed meta-data [16]. Prescriptions are usually made and revised by physicians regularly during clinical rounds, typically in the morning or early afternoon. Electronic decision support is generally immature and neither prescribing physicians nor dispensing nurses are warned if dose-adjustment be advised or even required.

We suspect that the need for dose-adjustment in patients with renal dysfunction often goes unrecognised. Thus, with this paper we study its predictability to inform clinicians and healthcare personnel upfront about which patients with renal dysfunction are at elevated risk of inappropriate drug dosing. To this end we used and compared predictive modelling methods from classical statistical modelling and machine learning.

## Methods

### Study design, patients and data

We conducted a register-based prediction study with prospective data for patients admitted to 12 public hospitals in two Danish regions comprising about 2.9 million persons (more than half the Danish population). We collected diagnosis data from the Danish National Patient Register, demographic data from the Danish Civil Registration System [17], as well as medication and biochemical data from electronic patient records. Diagnoses were encoded using the 10th revision of the International Classification of Diseases (ICD-10), drugs with the Anatomical and Therapeutic Chemical classification (ATC).

The units of analysis were inpatient admissions, defined as chains of successive in-hospital visits at most 24 hours apart. We included admissions starting between 1 October 2009 and 1 June 2016, with at least one eGFR measurement ≤30 during the first 24 hours of admission. We excluded minors (age <18 years). Admission time uses hour resolution (an admission starting at 9:54 is recorded as starting at 9:00) so to ensure at least 24 hours of observation time before inclusion, index was set at hour of admission + 25 hours. Prior sample-size estimation was foregone.

### Outcomes

The outcome variables were based on the daily rate = *r/E* of inappropriate doses during follow-up, capped at 30 days. *r* is the number of given inappropriate doses of select drugs cleared mainly renally and with narrow therapeutic indices; E the time-at-risk (figure 1). To obtain well-defined times-at-risk, we set the eGFR threshold to ≤30 ml/min/1.73m^2^ (unit omitted from here onward) and used the rules in supplementary table S1 for counting the number of inappropriate doses, based on the official reference guidelines for Danish physicians (pro.medicin.dk) as of January 2021.

**Figure 1:**
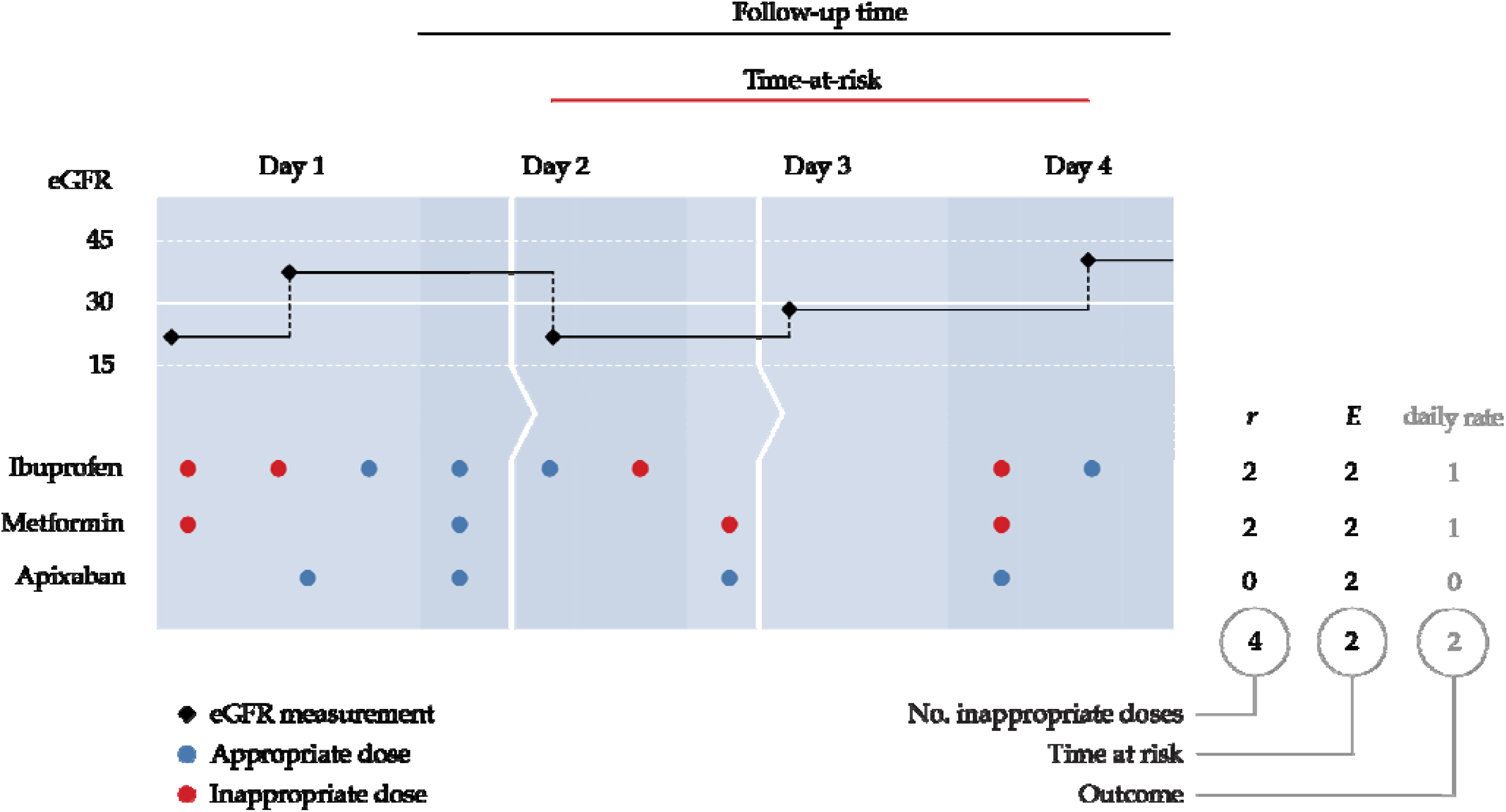
Deriving the outcome variables. This exemplary admission is composed of three successive in-patient visits (i.e. the patient has been transferred twice represented by the arrows). The admission is eligible because it spans more than 24 hours and an eGFR ≤30 was measured before index. Here, apixaban was given while the patient’s eGFR was ≤30, but dose reduction rendered these administrations appropriate.

We used two rules, one definitive (maximum daily dose = 0 mg) and one of dose-adjustment (reduced daily dose). Operationalisation of the definitive rule is straightforward: if the last eGFR ≤30, there should be no administrations until an eGFR >30 is measured. The dose-adjustment rule is slightly more involved as inappropriate dosing comes in two forms: (a) on a given day there are more than one eGFR measurements, of which at least one is ≤30, and the cumulative daily dose surpasses the threshold in the period(s) between above-threshold measurements, or (b) all eGFR measurements of a given day are ≤30 and the cumulative daily dose surpasses the threshold.

### Variables and features

Variables are original data (e.g. sex and age at admission) and features the results of rendering the variables appropriate as model inputs (e.g. one-hot-encoded day of admission). Based on clinical and pharmacological experience we hand-picked pertinent variables likely to be informative to the prediction problem and realistically available in the clinical setting. These fall into three categories. Demographic: age at admission (numeric), sex (binary). Clinical: number of distinct drugs (ATC level 5) administered between admission and index (numeric); therapeutic drug classes (ATC level 2) used between admission and index (one-hot-encoded); the Elixhauser score at admission (numeric, AQHR adaptation) [18]; ICD-10 chapters of diagnoses recorded in the past five years before admission (one-hot-encoded); record of chronic kidney failure in the past five years before admission (ICD-10 N18* diagnoses, one-hot-encoded). Contextual: hour of admission (numeric, transformed as *f(t) = abs*(12 − *t*); see supplementary figure S1); weekday of admission (one-hot-encoded); number of admissions in the past 5 years before admission (numeric).

Missing values, only present for hour of admission and discharge, were imputed by sampling from the empirical distributions of valid values.

### Models and training

We tried two model architectures (linear logistic ridge regression and artificial neural network) with several binary outcomes defined by increasing thresholds of the daily rate of inappropriate doses (>0, ≥1, ≥2, ≥3 and ≥5). The neural network models were multilayer perceptrons (MLPs) enabling speedy training and evaluation.

All admissions starting before 1 July 2015 were assigned to the development set (42,250 admissions [81%] of 27,253 patients) and the rest to the independent hold-out test set (10,201 admissions [19%] of 8,412 patients). Because admissions constitute the unit of analysis, some patients likely appear in both the development and test sets. Information may leak between the sets [19] so as a sensitivity analysis, we evaluated the performance also in the subset of test-set patients not in the development set.

We used the multivariate *TPEsampler* from *Optuna* [20] to find the best-performing hyperparameters by sampling 100 configurations, each using 5-fold stratified-and-grouped cross-validation, from the following proposal distributions (discrete values in round brackets, bounds of log-uniform distributions in squared): optimiser (Adam, RMSprop), learning rate [10^−6^, 10^−1^], activation function (tanh, sigmoid), L2 penalty [10^−6^, 10^−2^], number of hidden layers (1, 2, 3, 4), number of nodes per hidden layer [16, 32, 65, 128], batch size (32, 64, 128, 256, 512), class handling (see below).

Only relevant hyperparameters were sampled and we ran Optuna on linear and MLP models separately because they have disparate hyperparameter sets. MLP models with more hidden layers and more nodes therein can learn more complex relationships but become prone to overfitting which we countered with early stopping [21] and L2 regularisation (handles collinearity better than L1 regularisation) [22,23]. The batch size is the number of observations from which the model learns at a time; small batches can give outliers undue influence while full-batch training (batch size = number of units) can become computationally impractical [19]. Class imbalances in binary outcomes can misguide training, so we tested the following remedies: synthetic minority oversampling technique (SMOTE), random over-sampling of minority class, NearMiss, random under-sampling of majority class, class weighting, and none. SMOTE creates a dataset similar to the minority class but of the same size as the majority class [24]; NearMiss downsizes the majority class in a systematic way to retain as much information as possible in fewer data points [25]. Class weighting retains the original data but gives more weight to minority-class observations.

Hyperparameter optimisation models trained for maximum 500 epochs with 50-epoch patience on improvement in the validation loss. The final models were trained on the full development set until the loss reached that obtained in the best cross-validation fold for the best configuration [21].

### Evaluation and explanation

Discrimination was assessed with receiver operating characteristic (ROC) curves and areas under the ROC curves (AUROC), calibration-in-the-small by plotting decile-binned predicted probabilities against corresponding bin-wise observed event proportions [26] with 95% Jeffrey intervals [27]; results from a perfectly calibrated model fall on the diagonal. We used the decision-curve analytic framework to gauge the models’ potential clinical utility [28,29].

For explanation and scrutiny of prediction drivers, we used the SHAP DeepExplainer yielding one shap value per feature per unit [30]. The shap value for a risk prediction model is the absolute change in risk of a given unit’s value for each feature: the cohort-wide mean risk plus the sum of one unit’s shap values equals that unit’s risk.

### Analysis and ethics

The full analytical pipeline was built with Snakemake [31] (schematic overview in supplementary figure S2) to facilitate transparency and reproducibility; blinding was impractical and so foregone, but all analytic code is available online (DOI: <u>10.5281/zenodo.4560078</u>). Univariate distributions were summarised by median (inter-quartile range) and count (proportion), as appropriate. This report adheres to pertinent items in the MINIMAR guideline [32] and TRIPOD statement [33]. All data have been marshalled on Computerome, a secure high-performance Danish computing infrastructure, after obtaining approval from the Danish Patient Safety Authority (3-3013-1723; then competent authority for ethical approval), the Danish Data Protection Agency (DT SUND 2016-48, 2016-50, 2017-57) and the Danish Health Data Authority (FSEID 00003724).

## Results

Table 1 shows univariate summary statistics of the 52,451 admissions (42,250 + 10,201) of 35,665 patients (27,253 + 8,412) included in the study (see supplementary table S2 for extended version with all features). Patients in the test sets were similar to those in the development set with some notable exceptions. Fewer had received inappropriate doses, especially in the test-set patients not part of the development set who also had fewer previous admissions.

**Table 1:** Univariate summary statistics of select features. Values are median (inter-quartile range) and count (proportion) as appropriate. *Distinct patients* and *Distinct women* show counts of actual patients (as a patient can contribute more than one unit.)

| Variate | Development set<br>(N = 42,250) | Test set<br>(N = 10,201) | Test set<br>(not in devel. set)<br>(N = 5,980) |
| --- | --- | --- | --- |
| Women | 20,743 (49%) | 4,854 (48%) | 2,940 (49%) |
| Distinct patients | 27,253 | 8,412 | 5,341 |
| Distinct women | 13,759 (50%) | 4,049 (48%) | 2,629 (49%) |
| Time at risk, days | 3.5 (1.7–7.7) | 3.5 (1.7–7.2) | 2.9 (1.5–6.4) |
| Inappropriate doses (outcomes) |  |  |  |
| > 0 (at least one) | 3,786 (9.0%) | 1,080 (11%) | 740 (12%) |
| ≥ 1 daily | 2,241 (5.3%) | 588 (5.8%) | 333 (5.6%) |
| ≥ 2 daily | 1,236 (2.9%) | 288 (2.8%) | 108 (1.8%) |
| ≥ 3 daily | 783 (1.9%) | 171 (1.7%) | 56 (0.9%) |
| ≥ 5 daily | 366 (0.9%) | 64 (0.6%) | 9 (0.2%) |
| Admissions 5 years before admission |  |  |  |
| None | 4,988 (12%) | 1,082 (11%) | 1,074 (18%) |
| 1–2 | 10,100 (24%) | 2,367 (23%) | 1,873 (31%) |
| 3–4 | 7,712 (18%) | 1,919 (19%) | 1,232 (21%) |
| 5–6 | 5,490 (13%) | 1,303 (13%) | 685 (12%) |
| ≥ 7 | 13,960 (33%) | 3,530 (35%) | 1,116 (19%) |
| Drugs used between admission and index |  |  |  |
| None | 6,165 (15%) | 1,228 (12%) | 762 (13%) |
| 1–2 | 9,111 (22%) | 1,984 (19%) | 1,254 (21%) |
| 3–4 | 8,761 (21%) | 2,078 (20%) | 1,355 (23%) |
| 5–6 | 7,197 (17%) | 1,852 (18%) | 1,095 (18%) |
| ≥ 7 | 11,016 (26%) | 3,059 (30%) | 1,514 (25%) |
| Any diagnosis of chronic kidney failure | 13,470 (32%) | 3,391 (33%) | 732 (12%) |
| Top-5 ICD-10 chapters<sup>†</sup> |  |  |  |
| Cardiovascular (IX) | 25,757 (61%) | 6,392 (63%) | 3,283 (55%) |
| Genitourinary (XIV) | 23,025 (55%) | 5,819 (57%) | 2,306 (39%) |
| Lesions, external causes, etc. (XIX) | 20,275 (48%) | 4,749 (47%) | 2,481 (42%) |
| Metabolic-endocrine (IV) | 19,716 (47%) | 5,096 (50%) | 2,415 (40%) |
| Symptoms/abnormal findings (XVIII) | 18,663 (44%) | 5,711 (56%) | 2,882 (48%) |
| Top-5 drug classes<sup>‡</sup> |  |  |  |
| Analgesics (N02) | 15,740 (37%) | 4,367 (43%) | 2,506 (42%) |
| Systemic antibacterials (J01) | 14,719 (35%) | 3,257 (32%) | 1,938 (32%) |
| Diuretics (C03) | 13,966 (33%) | 3,672 (36%) | 1,951 (33%) |
| Antithrombotics (B01) | 11,842 (28%) | 3,181 (31%) | 1,795 (30%) |
| Antacids (A02) | 10,635 (25%) | 2,776 (27%) | 1,407 (24%) |

<sup>†</sup> ICD-10 chapters (Roman numbering) of diagnoses recorded in the last 5 years before admission.
<sup>‡</sup> Drug classes (ATC level 2) administered between admission and index.

In the development set, the median age was 77 years (IQR: 67-85) and 20,743 admissions (49%) were of 13,759 women (50%). The median time at risk was 3.5 days (inter-quartile range: 1.7–7.7) and at least one inappropriate dose was given in 3,786 admissions (9.0%); ≥1 inappropriate dose daily was given in 5.3% of admissions and ≥5 inappropriate doses daily were given in 0.9%. The target drugs most commonly given in inappropriate doses were ibuprofen (M01AE01, 4.1%) and metformin (A10BA02, 3.4%); inappropriate doses of the other target drugs were given in <1% of admissions.

Patients in 4,988 admissions (12%) had no admissions in the 5 years before inclusion; 13,960 (33%) had ≥7 previous admissions. The most common drug classes used between admission and index were analgesics (N02, 37%), systemic antibacterials (J01, 35%), diuretics (C03, 33%) antithrombotics (B01, 28%), and antacids (A02, 25%). Previous diagnoses were most commonly cardiovascular (chapter IX, 61%), genitourinary (XIV, 55%), related to i.a. lesions and external causes (XIX, 48%), endocrine-metabolic (IV, 47%), and symptoms/abnormal findings (XVIII, 44%).

Table 2 shows the hyperparameters of the best configurations with performance metrics of the final models (see also supplementary figures S3–S12). Generally, multi-layer perceptron (MLP) models performed slightly better than their linear counterparts, all obtaining AUROC’s between 0.77 and 0.81 in the test set (ROC curves in supplementary figures S13–S22). The MLP models more consistently showed good calibration in the development set. For daily rates >0, ≥1 and ≥2 both MLP and linear models were very well-calibrated in the test set (supplementary figures S23–S32). The decision curves did not suggest the clinical utility of the MLP models be superior to that of the linear (supplementary figures S33–S42).

**Table 2:** Performance metrics of final models and results of Optuna hyperparameter optimisation. AUROC: area under the receiver operating characteristic curve. MLP: multi-layer perceptron. Undersample: random sample of the size of the minority class, from the majority class. Oversample: randomly sample (with replacement) from the minority class until reaching a sample size equal to the size of the majority class. SMOTE: synthetic minority oversampling technique [24]. NearMiss: a method for non-random, systematic downsampling of the majority class while retaining as much information as possible [25].

| Parameter | Daily rate >0 |  | Daily rate ≥1 |  | Daily rate ≥2 |  | Daily rate ≥3 |  | Daily rate ≥5 |  |
| --- | --- | --- | --- | --- | --- | --- | --- | --- | --- | --- |
|  | Linear | MLP | Linear | MLP | Linear | MLP | Linear | MLP | Linear | MLP |
| AUROC |  |  |  |  |  |  |  |  |  |  |
| Development set | 0.80 | 0.81 | 0.81 | 0.83 | 0.81 | 0.84 | 0.82 | 0.83 | 0.82 | 0.83 |
| Test set | 0.77 | 0.79 | 0.78 | 0.79 | 0.79 | 0.79 | 0.81 | 0.81 | 0.78 | 0.80 |
| Test set (new patients) | 0.78 | 0.79 | 0.82 | 0.83 | 0.86 | 0.86 | 0.89 | 0.90 | 0.82 | 0.79 |
| Hyperparameters |  |  |  |  |  |  |  |  |  |  |
| Batch size | 512 | 128 | 512 | 32 | 32 | 64 | 256 | 256 | 64 | 64 |
| Class handling | Undersample | SMOTE | NearMiss | NearMiss | Oversample | SMOTE | Oversample | NearMiss | Oversample | None |
| L2 penalty | $1.28 \times 10^{-6}$ | $1.66 \times 10^{-6}$ | $3.02 \times 10^{-6}$ | $1.43 \times 10^{-6}$ | $4.38 \times 10^{-6}$ | $1.39 \times 10^{-6}$ | $1.43 \times 10^{-6}$ | $1.30 \times 10^{-6}$ | $1.09 \times 10^{-5}$ | $3.94 \times 10^{-6}$ |
| Learning rate | $1.79 \times 10^{-2}$ | $1.20 \times 10^{-4}$ | $1.92 \times 10^{-2}$ | $3.45 \times 10^{-4}$ | $6.73 \times 10^{-3}$ | $2.71 \times 10^{-4}$ | $3.76 \times 10^{-2}$ | $3.08 \times 10^{-4}$ | $2.11 \times 10^{-2}$ | $4.86 \times 10^{-4}$ |
| Optimiser | Adam | Adam | Adam | Adam | Adam | Adam | Adam | Adam | Adam | Adam |
| Activation function | — | tanh | — | sigmoid | — | tanh | — | sigmoid | — | sigmoid |
| No. hidden layers | — | 3 | — | 1 | — | 1 | — | 1 | — | 2 |
| Nodes per hidden layer | — | 8 | — | 8 | — | 32 | — | 32 | — | 8 |

The model-specific shap values offer some insights (supplementary figures S43– S53). First, many features contribute substantively to the predictions of daily rate >0 and ≥1 outcomes, while few features almost entirely drive the predictions for the other outcomes. Second, few features are the dominant prediction drivers across outcomes and models: use of anti-inflammatory, antirheumatic and antidiabetic drugs as well as diagnoses of chronic kidney failure. Third, sex and age contribute little to predictions. Fourth, using more distinct drugs (reflecting various levels of polypharmacy) pushes the risk up and using fewer drugs pulls the risk down. Fifth, the linear models tend to give most weight to relatively few features whereas the MLP models spread out the contributions across more features. Finally, the number of previous admissions (a proxy for frailty) became an increasingly important driver with increasing rarity of the outcome, in the MLP models.

Figure 2 shows the relationships between values of select features and their shap values and illustrates how MLP models capture highly non-linear effects and near-linear effects as appropriate (e.g. the effects of age at admission and number of previous admissions for daily rate >0.)

**Figure 2:**
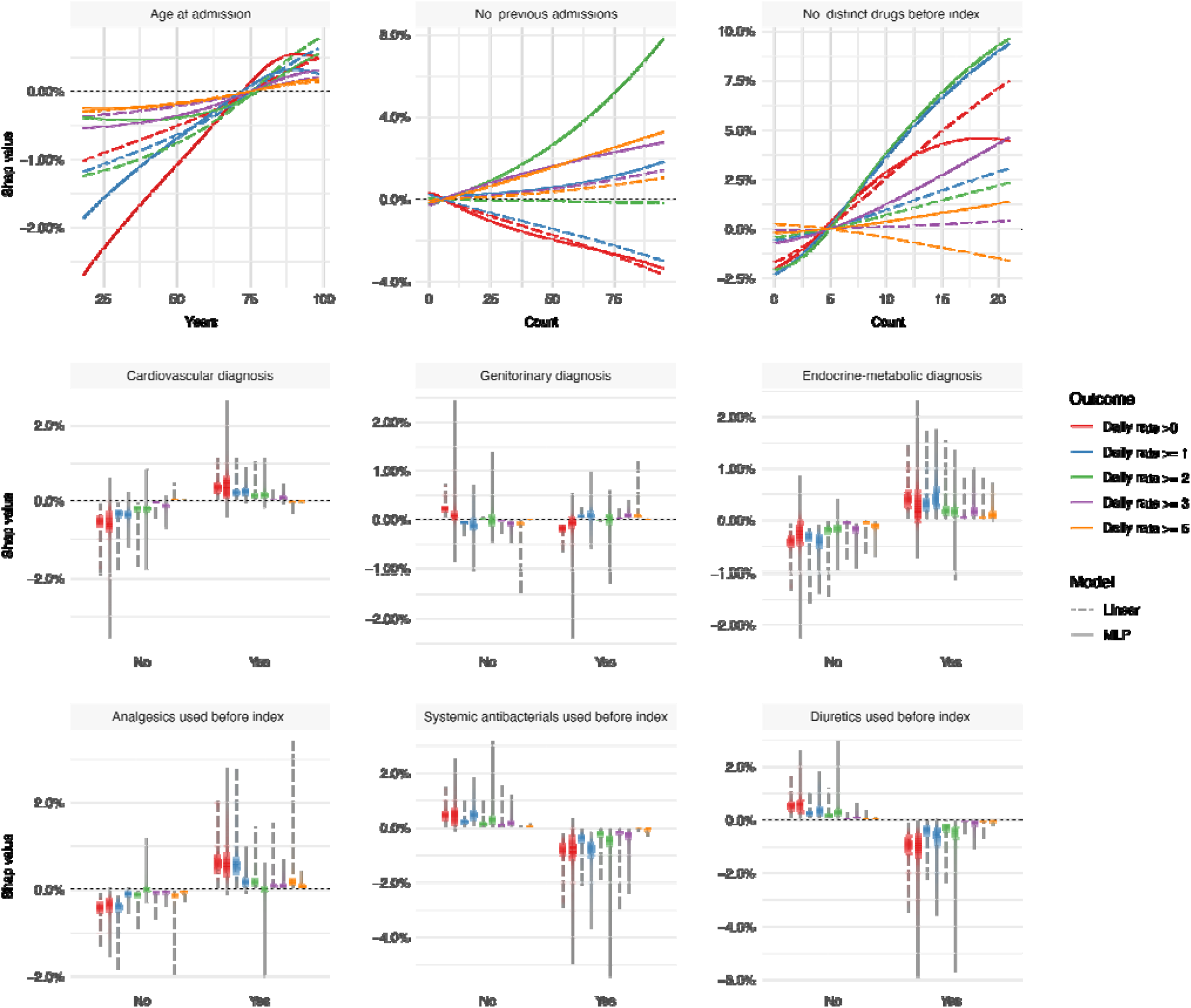
Bivariate relationships between values of select features (x axis) and their corresponding shap values (y axis). The continuous features are summarised by locally estimated scatterplot smoothing (LOESS), binary features by vertical density bands.

## Discussion

This study reveals that 9.0% of patients with reduced kidney function are exposed to inappropriate doses of selected renal risk drugs in the follow-up period. Our models performed quite well with AUROC’s between 0.77 and 0.81 with good calibration-in-the-small for daily rates >0 and ≥1, in the test set. For rarer outcomes (daily rates ≥2, ≥3 and ≥5) calibration suffered and clinical utility is unlikely to be substantive.

Apt intervention necessitates comprehension of the nature and extent of the problem. Use of renal risk drugs and associated problems, including inappropriate dosing, in patients with renal dysfunction is well-described [34-38]. A cross-sectional study of 83,000 American outpatient Veterans found that 32% of patients with creatinine clearance between 15 and 29 were given drugs at excessive doses considering their kidney function [39]. Medication burden had the strongest cooccurrence with inappropriate dosing and metformin was a prominent drug among those with inappropriate doses. This agrees with our findings although our study design has clearer temporality.

Some have called for a prediction tool to identify elderly at elevated risk of adverse drug reactions [40], a notion similar to ours in spirit but different in scope. Studies of factors associated with inadequate dose adjustment are few and often of retrospective nature eliciting relationships with characteristics after inappropriate doses have already been given. One study seeking to elicit factors associated with dosing appropriateness, using a logistic regression, reported the statistically strongest association to be with severity of chronic kidney failure (p-value = 7%) [41]. A similar study found dosing errors in 33% of the patients; *age* (odds ratio, OR: 1.05), *number of drug prescriptions* (OR: 1.1) and *number of drugs requiring dose adjustment* (OR: 2.0) were associated with dosing errors [42]. A third study found that, in patients with chronic kidney failure, *late-stage chronic kidney disease, number of prescribed drugs and presence of comorbidity* were associated with dosing errors. Ill-defined indices and times-at-risk render such enquiries of little use for a priori prediction and risk stratification: the ability to intervene presupposes a reliable estimate of risk in advance, before the event happens.

Carey et al. found only few factors to be genuinely predictive of potentially inappropriate prescribing in elderly outside the hospital setting [43]. Our models had AUROC’s (0.77–0.81) slightly higher than that of their model (0.76). In a prospective study from Norway [35] of internal-medicine patients with a mean age of 71 years, 35% received suboptimal doses; a composite variable (*number of clinical/pharmacological risk factors*) was quite strongly associated with non-optimal dosing (RR: 1.33), less so *number of drugs at admission* (RR: 1.09), whereas *sex* and *age* were not predictive of non-optimal dosing. Our results agree quite well with that finding, probably because the information captured by age and sex (essentially, proxies of comorbidity) is expressed explicitly in our feature set.

As such, our models fare quite well with performance metrics superior to those of other published models even though ours came from an independent and temporally distinct test set. Many studies employing machine learning models for predicting medical outcomes use normal split-sample validation, putting aside a random sample of the observations for testing. This has several logical and practical implications, perhaps most notably that a model developed with data collected between, say, 2005 and 2015 will likely perform better in a test case from 2013 than in one from 2017. The subset of our test set with patients not part of the development set is a conceptually appealing way to gauge how the model might perform in a new population. It does, however, distort the data and somewhat delink it from the clinical reality: some patients have previous admissions and those admitted for the first time are probably different from the rest.

### Strengths

Here we highlight five principal strengths of this study. First, this is by far the largest study of its kind to date. Second, time-series validation yielded realistic performance evaluation in distinct (future) data [44] vis-a-vis many articles on predictive modelling, perhaps most clearly seen in the surge of COVID-19 papers [45]. Third, our data were richer than in any other study in this area thanks to the combined diversity and reliability of longitudinal diagnostic data from the National Patient Register and deep phenotypic in-hospital data. Fourth, our summary statistics are well-aligned with descriptive studies of deviations from dosing recommendations, and the nature of the general patient population to which a model as ours would be applied [46]. Finally, the shap-value analysis suggests that the models picked up clinically relevant information without undue influence of individual predictors.

### Limitations

Like any study, this has potential limitations. First, albeit simple and elegant, using *only* eGFR as a proxy for kidney function is not always advisable [47]. It is, however, considered a reasonable metric for medicinal dosing [48] and used in Danish guidelines. Second, eGFR can be estimated in several ways [49] and both the 4-variable MRDR Study and CKD-EPI equations were used in our data. However, clinicians use the reported eGFR estimate as-is and both equations perform well for low eGFR values [50]. Third, hard thresholds on eGFR are arbitrary: the difference in kidney function between eGFRs of 29 and 31 is minuscule, but the cutoff must be set somewhere. Again, we stayed loyal to the guidelines as these are, nevertheless, what should support clinicians’ prescribing decisions. Fourth, many drugs have narrow and intermediate therapeutic indices. We focused on seven drugs cleared primarily by the kidneys and with narrow therapeutic indices that are fairly common in a Danish setting and span several important drug classes. The drugs included also allowed for reasonably harmonised rules of inappropriate dosing. Finally, our binary outcomes are soft endpoints and do constitute a simplification. Seemingly inappropriate doses could be conscious choices and the outcome variables do not capture information about actual toxicity experienced by the patient. However, the narrow therapeutic indices of the included drugs increase the likelihood of noxious effects without appropriate dose adjustment.

### Conclusion

Despite physicians’ awareness of the need for dose adjustment in patients with kidney dysfunction, a well-performing clinical decision support tool may help prevent such patients from “flying under the radar” in a busy clinical setting. Indeed, our models can flag patients at high risk of receiving >0 or ≥1 inappropriate dose daily.

A prospective evaluation is necessary to assess if these results transport to the clinic and if the models can offer genuine clinical utility for the patients. Receiving inappropriate doses is a soft endpoint, so clinical evaluation should consider also hard endpoints, either generic (e.g. length-of-stay, need for post-discharge rehabilitation and mortality) or specific ones related to the target drugs (e.g. transfusion and occurrence of known side-effects of these drugs).

## Data availability

Due to the sensitive nature of the data, we can neither offer access to nor share our data with third parties. Data can be obtained from the original sources upon request.

## Acknowledgements

The authors would like thank Innovation Fund Denmark (5153-00002B) and the Novo Nordisk Foundation (NNF14CC0001, NNF17OC0027594) for their financial contribution to BigTempHealth without which this study had not been possible. The funders played no role in designing, conducting, interpreting, or reporting this study.

## Contributions

Conceptualisation: BSKH, SEA. Data curation: BSKH, CLR. Formal analysis: BSKH. Methodology: APN, BSKH, DP, HCTM, ND. Software: BSKH. Code review: CLR, DP, HCTM. Drafting: BSKH. Funding acquisition: SB, SEA. Resources: SB, SEA. Supervision: SEA. Review: All.

## Conflicts of interest

The authors declare the following competing interests:

- BSKH: None
- CRL: None
- DP: None
- HCTM: None
- ND: None
- APN: None
- SB reports ownerships in Intomics A/S, Hoba Therapeutics Aps, Novo Nordisk A/S, Lundbeck A/S, and managing board memberships in Proscion A/S and Intomics A/S outside the submitted work
- SEA: None

## Notes

### Funding Statement

This work was supported by the Novo Nordisk Foundation (grants NNF14CC0001 and NNF17OC0027594) and the Danish Innovation Fund (grant 5153-00002B).

### Author Declarations

Approval was obtained from the Danish Patient Safety Authority (3-3013-1723; then competent authority for ethical approval), the Danish Data Protection Agency (DT SUND 2016-48, 2016-50, 2017-57) and the Danish Health Data Authority (FSEID 00003724).

