## Supplement for "Identifying patients at high risk of inappropriate drug dosing in periods with renal dysfunction": Supplement.html

Development of a prediction model to identify patients at high risk of inappropriate drug dosing in periods with renal dysfunction


### Development of a prediction model to identify patients at high risk of inappropriate drug dosing in periods with renal dysfunction

##### Supplement

###### Benjamin Skov Kaas-Hansen

###### 19 Feburary 2021

- Supplementary tables
  - Table S1: Maximum doses in periods when eGFR is as indicated. ATC codes in brackets.
  - Table S2 (extended version of table 1): Univariate summary statistics of the three data sets. Values are median (inter-quartile range) and N (%).
- Supplementary figures
  - Figure S1: Mapping of admission hour, two alternatives.
  - Figure S2: The so-called rulegraph of the our Snakemake pipeline illustrating the end-to-end workflow with dependencies between processing, training and visualisation steps.
- Optuna hyperparameter optimisation
  - Figure S3: MLP, daily rate > 0.
  - Figure S4: Linear, daily rate > 0.
  - Figure S5: MLP, daily rate >= 1.
  - Figure S6: Linear, daily rate >= 1.
  - Figure S7: MLP, daily rate >= 2.
  - Figure S8: Linear, daily rate >= 2.
  - Figure S9: MLP, daily rate >= 3.
  - Figure S10: Linear, daily rate >= 3.
  - Figure S11: MLP, daily rate >= 5.
  - Figure S12: Linear, daily rate >= 5.
- ROC curves
  - Figure S13: MLP, daily rate > 0.
  - Figure S14: Linear, daily rate > 0.
  - Figure S15: MLP, daily rate >= 1.
  - Figure S16: Linear, daily rate >= 1.
  - Figure S17: MLP, daily rate >= 2.
  - Figure S18: Linear, daily rate >= 2.
  - Figure S19: MLP, daily rate >= 3.
  - Figure S20: Linear, daily rate >= 3.
  - Figure S21: MLP, daily rate >= 5.
  - Figure S22: Linear, daily rate >= 5.
- Calibration plots
  - Figure S23: MLP, daily rate > 0.
  - Figure S24: Linear, daily rate > 0.
  - Figure S25: MLP, daily rate >= 1.
  - Figure S26: Linear, daily rate >= 1.
  - Figure S27: MLP, daily rate >= 2.
  - Figure S28: Linear, daily rate >= 2.
  - Figure S29: MLP, daily rate >= 3.
  - Figure S30: Linear, daily rate >= 3.
  - Figure S31: MLP, daily rate >= 5.
  - Figure S32: Linear, daily rate >= 5.
- Decision curves
  - Figure S33: MLP, daily rate > 0.
  - Figure S34: Linear, daily rate > 0.
  - Figure S35: MLP, daily rate >= 1.
  - Figure S36: Linear, daily rate >= 1.
  - Figure S37: MLP, daily rate >= 2.
  - Figure S38: Linear, daily rate >= 2.
  - Figure S39: MLP, daily rate >= 3.
  - Figure S40: Linear, daily rate >= 3.
  - Figure S41: MLP, daily rate >= 5.
  - Figure S42: Linear, daily rate >= 5.
- SHAP plots
  - Figure S43: Summary plot of shap values across studies
  - Figure S44: MLP, daily rate > 0
  - Figure S45: Linear, daily rate > 0
  - Figure S46: MLP, daily rate >= 1
  - Figure S47: Linear, daily rate >= 1
  - Figure S48: MLP, daily rate >= 2
  - Figure S49: Linear, daily rate >= 2
  - Figure S50: MLP, daily rate >= 3
  - Figure S51: Linear, daily rate >= 3
  - Figure S52: MLP, daily rate >= 5
  - Figure S53: Linear, daily rate >= 5

---

This document contains all supplementary tables and figures for the above-mentioned paper. The figures within each domain all share the same caption, and so to avoid cluttering captions are given only once, under the domain heading.

---

###### Supplementary tables

###### Table S1: Maximum doses in periods when eGFR is as indicated. ATC codes in brackets.

###### Table S2 (extended version of table 1): Univariate summary statistics of the three data sets. Values are median (inter-quartile range) and N (%).

###### Supplementary figures

###### Figure S1: Mapping of admission hour, two alternatives.

Top: Mapping of admission hour (x axis) to unit circle (blue) and distance from midday (red). Bottom: The unit-circle mapping loses the interval nature of the admission hour on the original scale, whereas the distance-to-midday maintains this quality.

###### Figure S2: The so-called rulegraph of the our Snakemake pipeline illustrating the end-to-end workflow with dependencies between processing, training and visualisation steps.

###### Optuna hyperparameter optimisation

Sampled hyperparameter configurations. Point colours represent the loss value: blue = low (preferable), red = high, grey = above the third quartile. Dashed lines indicate the best configuration.

###### Figure S3: MLP, daily rate > 0.

###### Figure S4: Linear, daily rate > 0.

###### Figure S5: MLP, daily rate >= 1.

###### Figure S6: Linear, daily rate >= 1.

###### Figure S7: MLP, daily rate >= 2.

###### Figure S8: Linear, daily rate >= 2.

###### Figure S9: MLP, daily rate >= 3.

###### Figure S10: Linear, daily rate >= 3.

###### Figure S11: MLP, daily rate >= 5.

###### Figure S12: Linear, daily rate >= 5.

###### ROC curves

Receiver-operating characteristic (ROC) curves in the development and test sets.

###### Figure S13: MLP, daily rate > 0.

###### Figure S14: Linear, daily rate > 0.

###### Figure S15: MLP, daily rate >= 1.

###### Figure S16: Linear, daily rate >= 1.

###### Figure S17: MLP, daily rate >= 2.

###### Figure S18: Linear, daily rate >= 2.

###### Figure S19: MLP, daily rate >= 3.

###### Figure S20: Linear, daily rate >= 3.

###### Figure S21: MLP, daily rate >= 5.

###### Figure S22: Linear, daily rate >= 5.

###### Calibration plots

Calibration curves in the development and test sets.

###### Figure S23: MLP, daily rate > 0.

###### Figure S24: Linear, daily rate > 0.

###### Figure S25: MLP, daily rate >= 1.

###### Figure S26: Linear, daily rate >= 1.

###### Figure S27: MLP, daily rate >= 2.

###### Figure S28: Linear, daily rate >= 2.

###### Figure S29: MLP, daily rate >= 3.

###### Figure S30: Linear, daily rate >= 3.

###### Figure S31: MLP, daily rate >= 5.

###### Figure S32: Linear, daily rate >= 5.

###### Decision curves

Decision curves based on the test set. The curves show the clinical utility (in the unit of standardised net benefit) of intervening in all patients (magenta), no patients (dark grey, follows the x axis) and patients identified by the model (blue).

###### Figure S33: MLP, daily rate > 0.

###### Figure S34: Linear, daily rate > 0.

###### Figure S35: MLP, daily rate >= 1.

###### Figure S36: Linear, daily rate >= 1.

###### Figure S37: MLP, daily rate >= 2.

###### Figure S38: Linear, daily rate >= 2.

###### Figure S39: MLP, daily rate >= 3.

###### Figure S40: Linear, daily rate >= 3.

###### Figure S41: MLP, daily rate >= 5.

###### Figure S42: Linear, daily rate >= 5.

###### SHAP plots

The individual shap plots (figures S44-53) below all visualise the the distributions as density bands of shap values by feature. Blue represents low feature values (0 for binary features) and red high feature values (1 for binary features). Continuous features were binned into deciles.

###### Figure S43: Summary plot of shap values across studies

Feature values were binned into at most 5 bins, and each bins is represented by one point: the x axis is the mean shap value for each bin, the colour illustrates the spectrum of feature values (blue = low, red = high). Points are connected by lines to aid reading, solid and dashes lines represent MLP and linear models.

###### Figure S44: MLP, daily rate > 0

###### Figure S45: Linear, daily rate > 0

###### Figure S46: MLP, daily rate >= 1

###### Figure S47: Linear, daily rate >= 1

###### Figure S48: MLP, daily rate >= 2

###### Figure S49: Linear, daily rate >= 2

###### Figure S50: MLP, daily rate >= 3

###### Figure S51: Linear, daily rate >= 3

###### Figure S52: MLP, daily rate >= 5

###### Figure S53: Linear, daily rate >= 5
